# Robust Feature Engineering Techniques for Designing Efficient Motor Imagery-Based Systems Using Multidomain Electroencephalography

**DOI:** 10.64898/2026.11.11.26367815

**Authors:** Syed Saim Ahmed, Soyiba Jawed, Mahnoor Ahmed Khan, Zoha Zahid Fazal, Tehmina Shehryar, Syeda Muneeba Bukhari, Rizwan Ahmed Khan

## Abstract

Motor imagery-based brain-computer interfaces (BCls) have potential applications in neurorehabilitation; however, the high dimensionality and complexity of electroencephalography (EEG) signals present challenges for accurate classification. This study aimed to improve the classification of motor and motor imagery activities using multidomain feature engineering and feature selection on the MILimbEEG dataset. Time-domain, frequency-domain, and wavelet-based features were extracted from 16-channel EEG signals, yielding 416 features. Maximum relevance minimum redundancy (MRMR) was then applied to select the four most informative and minimally redundant features. These features were used to train support vector machine (SVM), K-nearest neighbors (KNN), Na"ive Bayes, and decision tree classifiers, with performance evaluated using 10-fold cross-validation based on testing accuracy, precision, recall, and Fl-score. The Gaussian-kernel SVM achieved the highest testing accuracies of 92.50% for motor activities and 95.48% for motor imagery activities. For the highest-performing motor imagery classification, precision, recall, and Fl-score were 86.97%, 88.33%, and 87.04%, respectively. These findings demonstrate that reducing 416 multidomain EEG features to four discriminative features using MRMR can maintain strong classification performance. The proposed feature-engineering framework may support the development of computationally efficient EEG-based BCI systems for neurorehabilitation applications.

**Highlights:**

- Multidomain feature engineering extracts 416 features from 16-channel EEG signals.
- MRMR reduces 416 EEG features to four discriminative features for classification.
- Gaussian SVM achieves 95.48% accuracy for motor imagery classification.
- Gaussian SVM achieves 92.50% accuracy for motor activity classification.

## Introduction

Motor disabilities substantially affect independence and quality of life, highlighting the importance of rehabilitation technologies1. Brain-computer interfaces (BCls) enable communication and control of assistive devices by translating neural activity into external commands^2^. Among these approaches, non-invasive electroencephalography (EEG)-based BCls are particularly promising for neurorehabilitation because of their accessibility, portability, and relatively low cost3. Motor imagery (Ml)-based BCls are especially relevant because they use neural activity associated with imagined movements without requiring overt motor execution, offering potential applications for individuals with impaired voluntary movement^4^.

Despite these advantages, reliable Ml classification remains challenging because EEG signals are non-stationary, have low signal-to-noise ratios, and exhibit substantial inter-individual variability5. Accurate decoding therefore depends on extracting discriminative signal characteristics and selecting informative features while minimizing redundancy. Machine learning (ML) approaches have been widely investigated to address these challenges, including conventional classifiers and deep learning methods^6,7^. The general ML workflow, encompassing feature extraction, model training, and prediction, is illustrated in **Figure 1**. However, high-dimensional feature representations, limited training data, and variability across individuals continue to affect classification performance and generalizability^5,8^.

**Figure 1.**
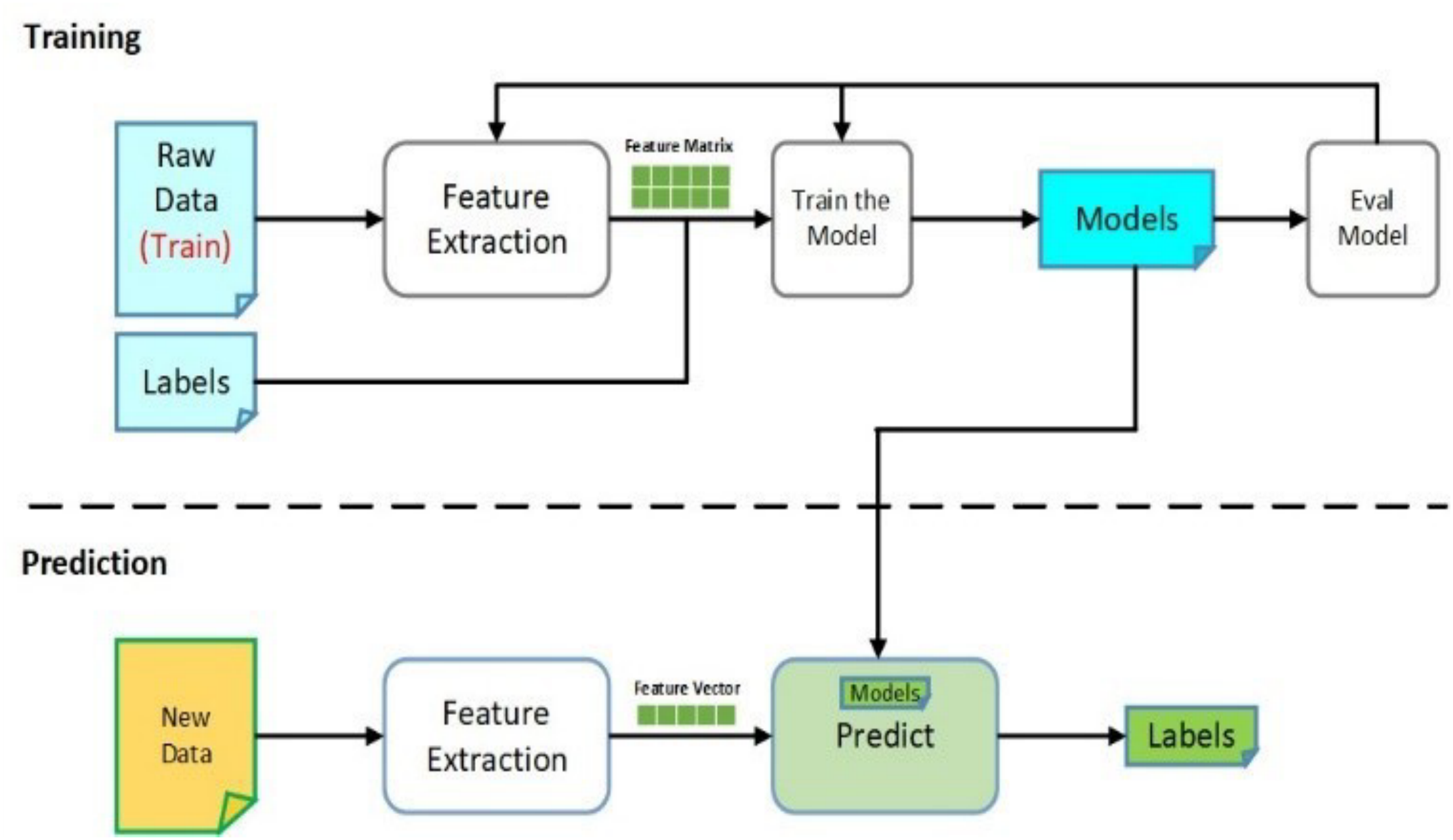
Overview of the machine learning workflow for EEG-based classification. Abbreviations: EEG, electroencephalography.

Previous studies have investigated time-domain, frequency-domain, and time-frequency approaches to improve Ml classification. Feature-selection techniques, including mutual information-based methods, have been employed to identify discriminative EEG characteristics and reduce feature dimensionality9. Although these strategies have demonstrated promising results, the performance of EEG classification systems remains dependent on dataset characteristics, feature representation, and validation methodology^7,8^. Consequently, identifying compact feature sets that retain relevant information remains an important objective in EEG signal processing.

The availability and characteristics of EEG datasets also influence the development and evaluation of Ml-based BCls. Widely used datasets from BCI Competition IV contain recordings from relatively small participant groups, including nine participants in Dataset 2a and nine in Dataset 2b^10^. Such datasets have contributed substantially to algorithm development but may limit the assessment of performance across diverse individuals. Reviews of publicly available motor imagery and execution datasets have emphasized the importance of dataset selection and standardized evaluation when comparing classification approaches^11^.

The MILimbEEG dataset^12^ provides recordings of upper-and lower-limb motor execution and imagery tasks, enabling investigation of multiple movement classes using non-invasive EEG^13^. Its inclusion of both motor execution and imagery conditions provides an opportunity to examine how feature representations and classification algorithms perform across different movement comparisons. However, further investigation is needed to determine whether multidomain feature extraction combined with feature selection can support accurate classification using a substantially reduced feature set.

Accordingly, the present study investigates a multidomain feature-engineering framework for classifying motor execution and imagery signals from the MILimbEEG dataset. Time-domain, frequency-domain, and wavelet-based features are extracted from 16-channel EEG recordings, followed by MRMR feature selection to reduce dimensionality. The selected features are evaluated using support vector machine (SVM), K-nearest neighbors (KNN), Na"ive Bayes, and decision tree classifiers, with performance assessed through testing accuracy, precision, recall, and Fl-score. The principal contribution is the evaluation of a compact feature representation derived from complementary EEG signal domains for motor execution and imagery classification.

## Methodology

### Dataset Description

The MllimbEEG dataset contains EEG recordings acquired during upper-and lower-limb motor execution and imagery tasks^13^. The original dataset includes 60 participants performing six movement tasks: closing the left hand (CLH), closing the right hand (CRH), dorsiflexion of the left foot (DLF), plantar flexion of the left foot (PLF), dorsiflexion of the right foot (DRF), and plantar flexion of the right foot (PRF), along with resting conditions as shown in **Figure 2**.

**Figure 2.**
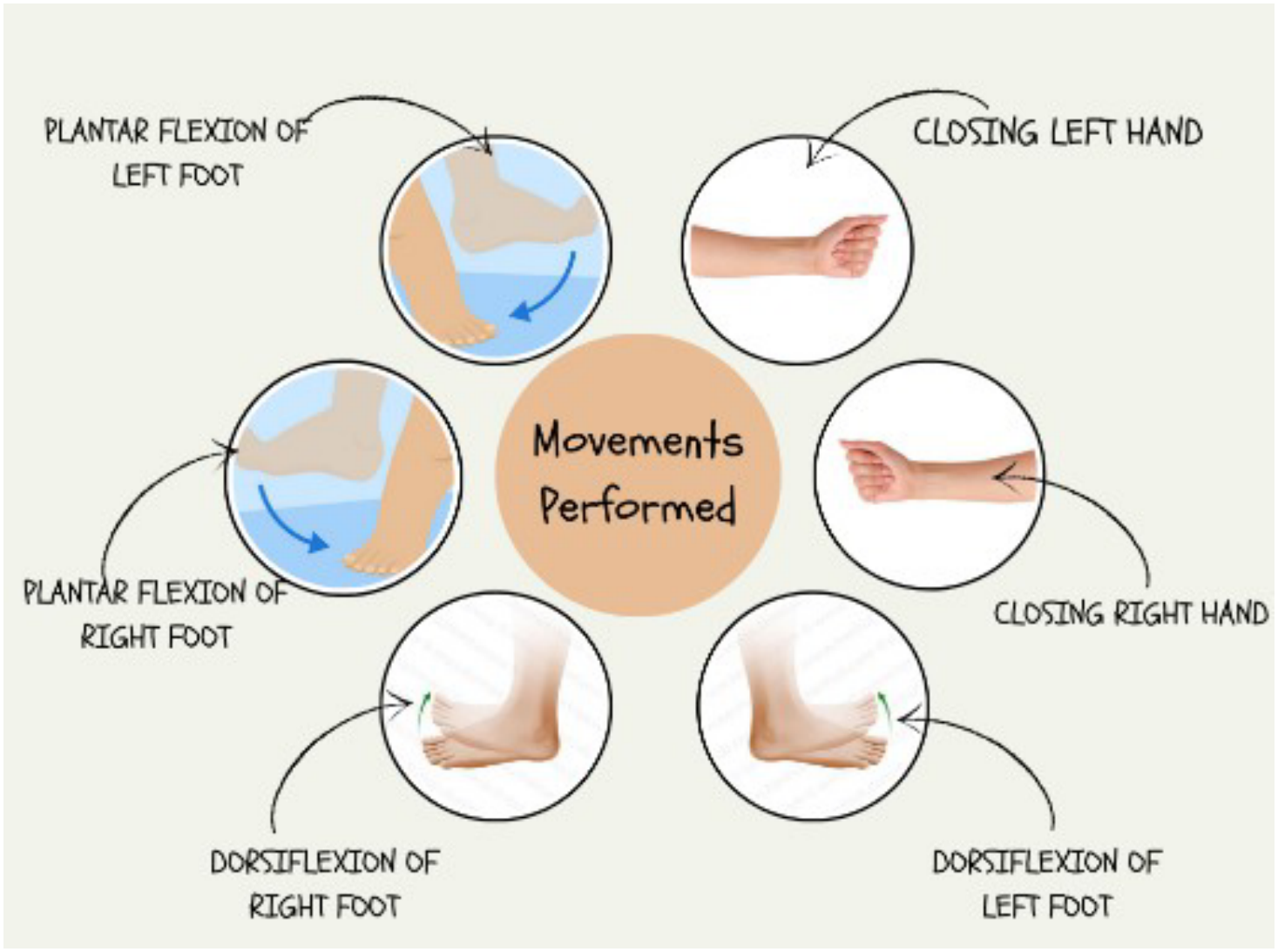
Upper-and lower-limb movements performed during motor execution and motor imagery tasks.

EEG signals were recorded using a 16-channel OpenBCI Cyton + Daisy system at a sampling frequency of 125 Hz, with electrodes positioned according to the international 10/10 system, as illustrated in **Figure 3**. Recordings were stored as CSV files containing 500 samples per channel. The present study used recordings from 24 participants, comprising 2,976 available files. The overall analytical framework is shown in **Figure 4**.

**Figure 3.**
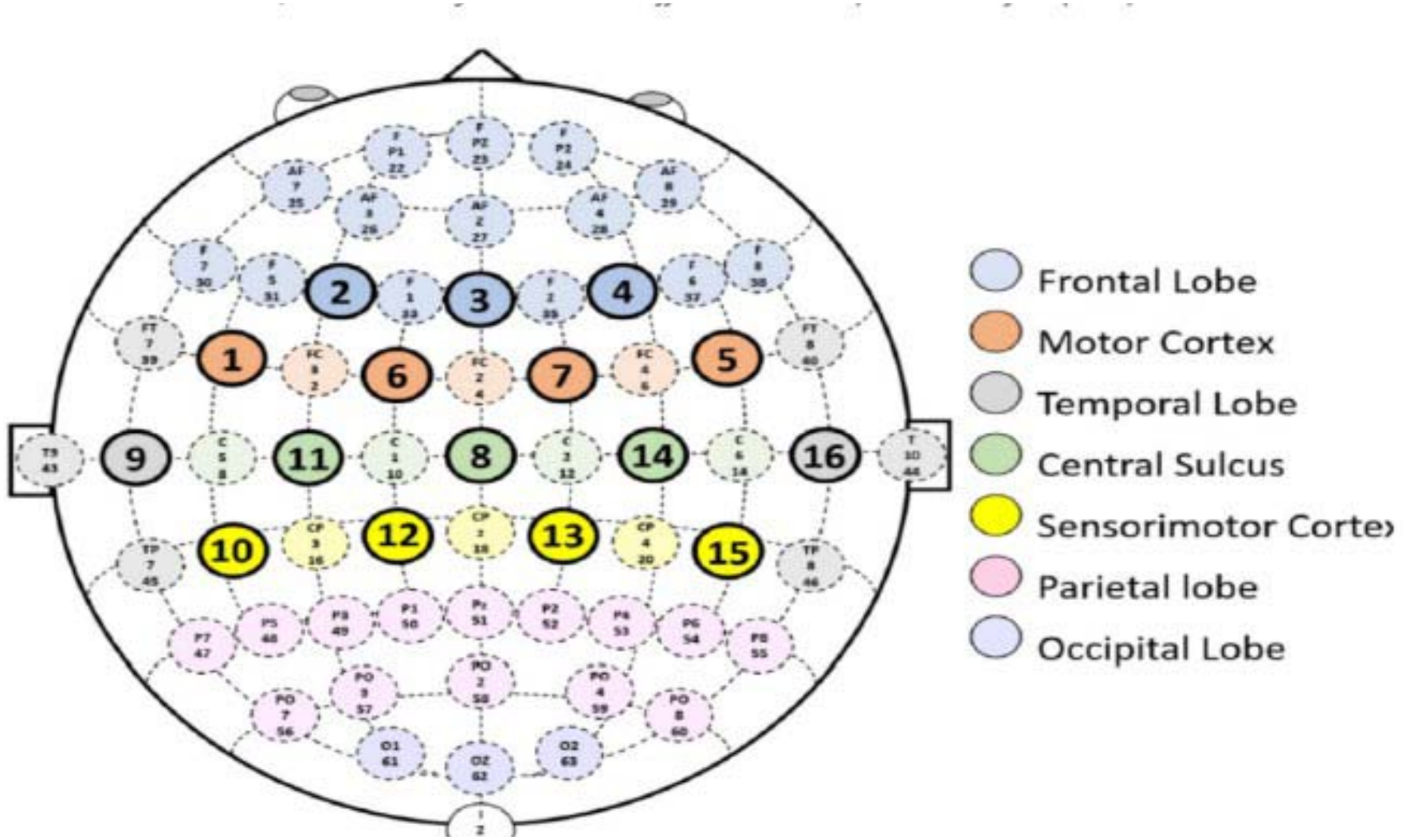
Electrode placement for EEG signal acquisition. Abbreviations: EEG, eIectroeneephalography.

**Figure 4.**
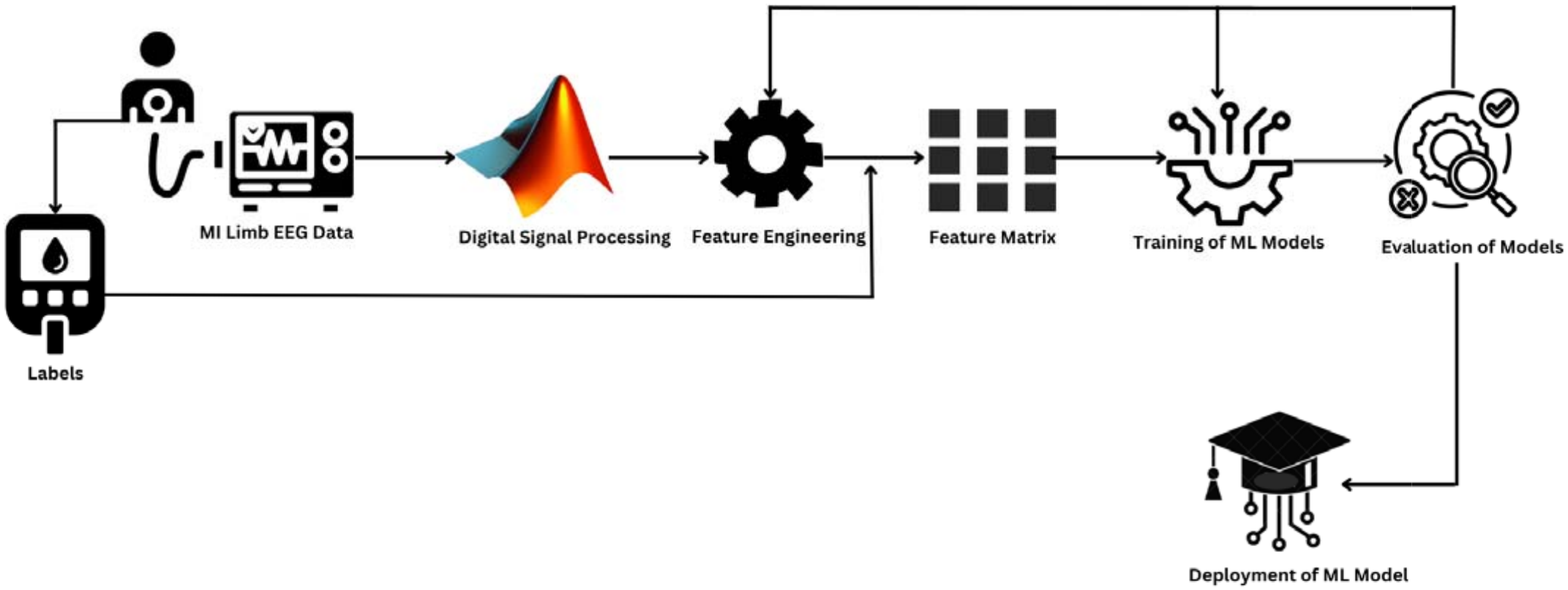
Proposed multidomain feature-engineering and classification workflow. Abbreviations: EEG, electroencephalography; ML, machine learning.

### Preprocessing and Feature Extraction

Of the 2,976 available recordings, 384 files (16 recordings per participant) were selected for analysis. EEG signals were normalized by dividing each channel’s values by its maximum amplitude. No additional digital filtering was applied.

A total of 416 features were extracted from the 16 EEG channels, comprising 160 time-domain, 96 frequency-domain, and 160 wavelet-based features. Ten time-domain features were calculated per channel, as summarized in **Table 1**.

**Table 1.** Time-domain features extracted from electroencephalography signals. Abbreviations: EEG, electroencephalography; HA, Hjorth activity; HC, Hjorth complexity; HM, Hjorth mobility; ME, mean energy; MTE, mean Teager’s energy; NFD, normalized first difference; NSD, normalized second difference; SD, standard deviation; ShEn, Shannon entropy.

| **Feature Name** | **Short Summary** |
|---|---|
| Mean | Mean Amplitude Value of the EEG signal. V. Raj et al. identified mean as a feature capable of distinguishing attention-demanding tasks from resting states35. |
| Standard Deviation (SD) | SD is the square root of variance. It is an effective parameter for comparing real and imagery motion patterns. |
| Mean Energy (ME) | The power of a signal to identify a series in the power curve at any given time is represented by its ME. It provides physiologically rational and empirically effective information about the EEG signals. |
| Mean Teager's Energy (MTE) | MTE calculates the difference between the squared of the first derivative and the product of the signal and its second derivative. It is used to measure the variation between successive samples of digital signals, and thus it helps to estimate a signal's instantaneous energy. |
| Normalized First Difference (NFD) | The first difference in the normalized EEG signal is the correlation between the current and prior normalized EEG signal data. The NFD helps us to see how quickly or slowly brain activity is changing. |
| Normalized Second Difference (NSD) | NSD is a measure of how the first difference (the change in signal from one point to the next) changes over time. It is used for analyzing how the speed of brain activity changes. |
| Shannon Entropy (ShEn) | A measurement of the uncertainty (or variability) connected to random variables is called ShEn, or simply entropy. It helps in analyzing the complexity and variability of brain activity |
| Hjorth Activity (HA) | HA is a time-domain parameter that measures the mean power or activity of a signal. A higher activity value shows more brain activity, while a lower activity value indicates less. |
| Hjorth Complexity (HC) | HC gives an estimate of a signal's bandwidth and reveals how closely a signal resembles a pure sinusoidal waveform. This measures the signal's complexity in terms of how many different frequencies are present. A higher complexity value suggests a more intricate signal with diverse frequency components, while a lower complexity indicates a simpler signal. |
| Hjorth Mobility (HM) | The percentage of SD or power spectrum's mean frequency is HM. It can be expressed as the square root of the signal's first derivative's activity divided by the signal's activity. Higher mobility means the signal is changing quickly, while lower mobility indicates more stability. |

Frequency-domain characteristics were extracted using Welch’s power spectral density (PSD) estimation method, which averages modified periodograms from overlapping signal segments**^14^**:

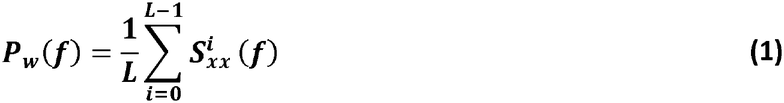

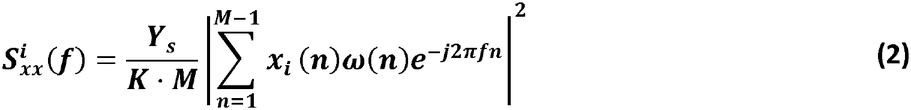

where Si (f) is the optimized periodogram, (f = ts) is the normalized frequency variable, (Y s) is the scaling factor, w(n) is windowing function, L is the length of the mark and K is the normalized constant defined as:

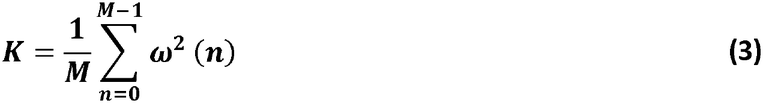

A Hanning window was used for PSD estimation. Frequency-domain features included average amplitude and band power across the delta, theta, alpha, beta, and gamma bands.

Time-frequency features were extracted using the discrete wavelet transform (DWT), which decomposes non-stationary EEG signals into approximation and detail coefficients^15^. The decomposition is expressed as:

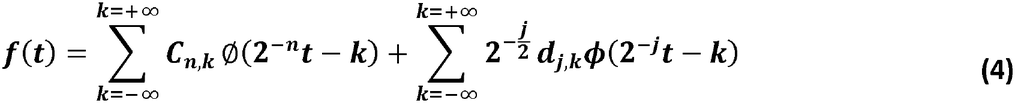

where dJ, k) and C(n, k) represent the approximation and detail coefficients, respectively, n is the level, and cj) is the function of scale. The Daubechies 4 (db4) wavelet was used to extract wavelet-based characteristics from each channel.

### Case Preparation

The extracted features were organized across 13 task conditions, comprising baseline, six motor execution tasks, and six corresponding motor imagery tasks. Each recording was represented by 416 features: 160 time-domain, 96 frequency-domain, and 160 wavelet-based features. The resulting feature matrix was organized into 11 binary classification cases, as defined in **Table 2**.

**Table 2.** Classification cases for motor execution and motor imagery tasks relative to the resting baseline. Abbreviations: BEO, both eyes open; CLH, closing left hand; CRH, closing right hand; DLF, dorsiflexion of left foot; ORF, dorsiflexion of right foot; PLF, plantar flexion of left foot; PRF, plantar flexion of right foot.

| **Case** | **Motor Activity Group** | **Comparison Condition** |
|---|---|---|
| 1 | CLH, CRH, DLF, PLF, DRF, PRF | BEO |
| 2 | CLH or CRH | BEO |
| 3 | DLF, PLF, DRF, PRF | BEO |
| 4 | CRH | BEO |
| 5 | CLH (Motor/Motor Imagery) | BEO |
| 6 | DRF and PRF | BEO |
| 7 | DRF | BEO |
| 8 | PRF | BEO |
| 9 | DLF, PLF | BEO |
| 10 | DLF | BEO |
| 11 | PLF | BEO |

### Feature Selection

To reduce feature dimensionality, PCA was initially evaluated. Although three principal components retained approximately 95% of the variance, they did not provide satisfactory class separation. Consequently, MRMR feature selection was employed to identify informative features while minimizing redundancy.

The MRMR criterion is defined as:

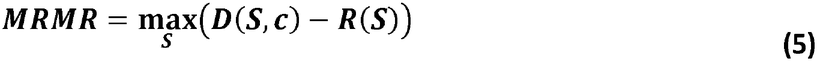

The equation shows optimal features can be selected with maximal relevance D(S, c) and minimal redundancy R(S) using MRMR.

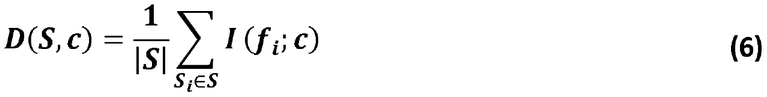

Here, |S| is the number of features in sample feature set S = {f1, f2, …, f_n_} and l(f_i_; c) is the mutual information between the feature f_i_ and the class c. The mutual information l(x; y) is defined mathematically as:

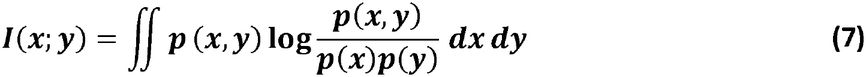

where Sp(x, y) is the joint probability density of random variables x and y, and Sp(x) and Sp(y) are the marginal probability densities of x and y, respectively. The redundancy of all features in Sis the mean of all mutual information between feature fi and feature f_j_.

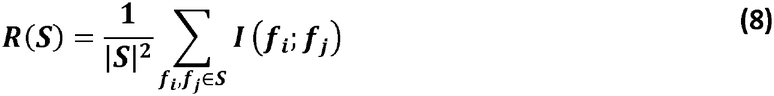

MRMR reduced the original 416-dimensional feature space to four selected features. **Figure 5** illustrates the separation obtained using three of the highest-ranked features for Case I.

**Figure 5.**
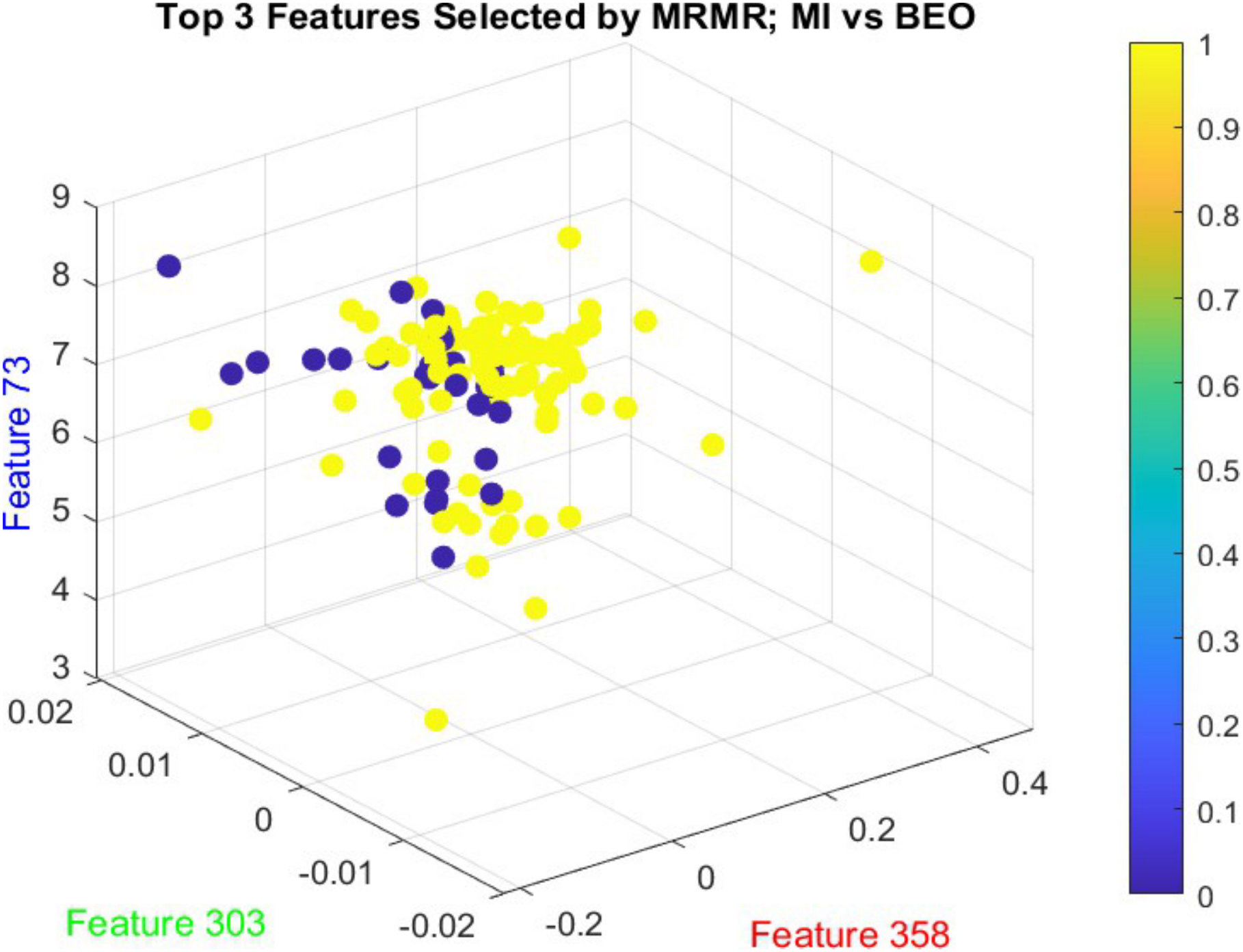
Three-dimensional visualization of features selected using maximum relevance minimum redundancy. Abbreviations: BEO, both eyes open; Ml, motor imagery; MRMR, maximum relevance minimum redundancy.

### Training Classification

The selected features were evaluated using Gaussian- and polynomial-kernel SVM, KNN with Euclidean and Mahalanobis distances, Na"ive Bayes, and decision tree classifiers.

For KNN, the number of neighbors was determined usingthe expression:

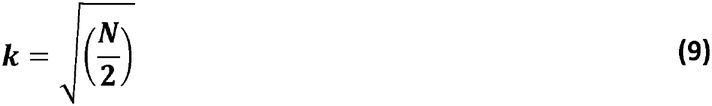

Where N is the total number of samples^16^.

### Performance Evaluation

Model performance was evaluated using 10-fold cross-validation^17^. The dataset was divided into ten subsets, with nine used for training and one for testing in each iteration. Performance was summarized using average training accuracy, testing accuracy, precision, recall, and Fl-score.

## Results

The classification performance of the evaluated models for motor imagery and motor execution is presented in **Tables 3 and 4**, respectively. Following multidomain feature extraction, MRMR reduced the initial 416 features to four selected features. Performance was evaluated using 10-fold cross-validation and summarized using training accuracy, testing accuracy, precision, recall, and Fl-score. Across the 11 classification cases defined in **Table 2**, Gaussian-kernel SVM achieved the highest testing accuracies for the combined movement comparisons (Case I), reaching 95.48% for motor imagery and 92.50% for motor execution.

**Table 3.**
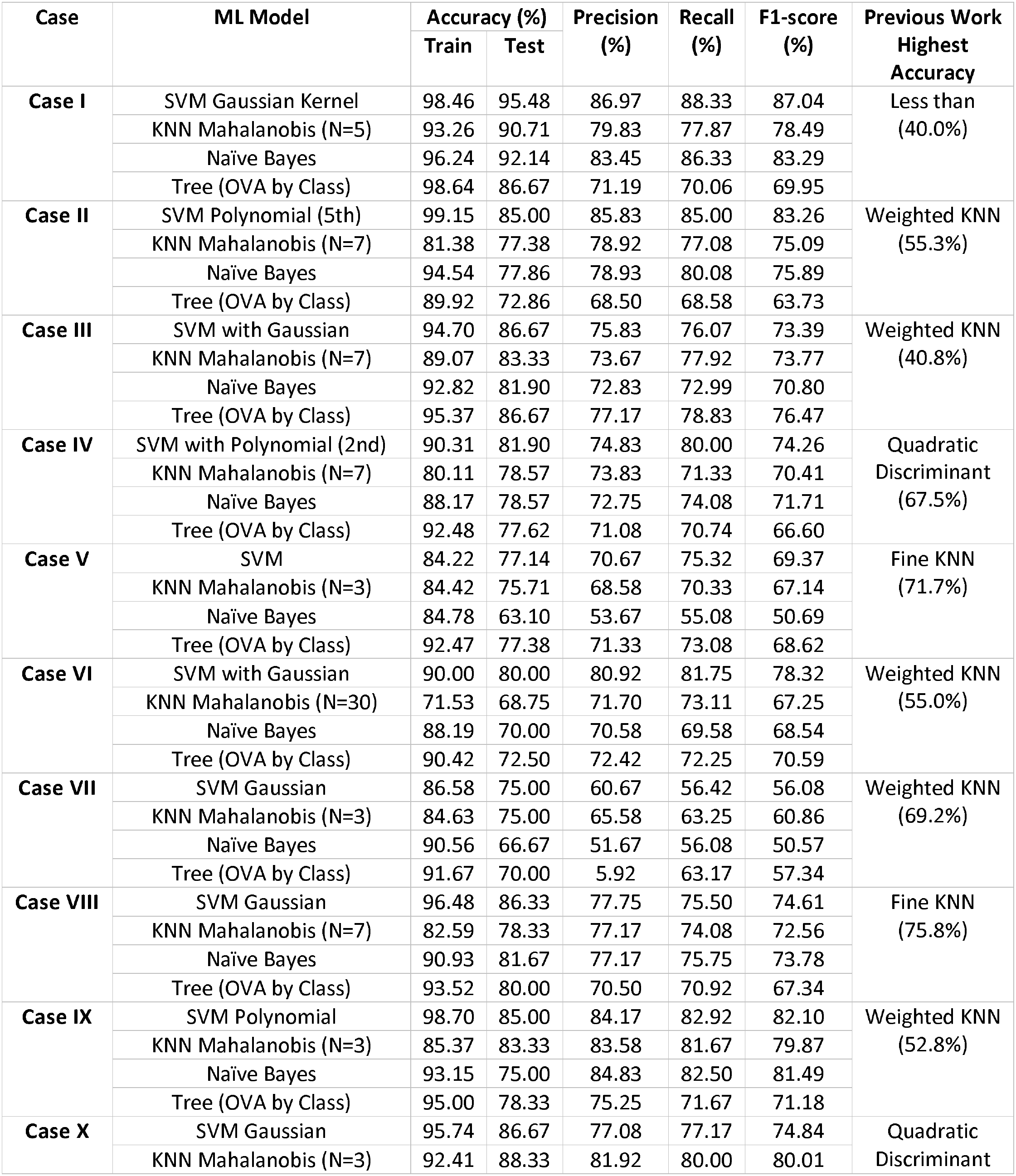
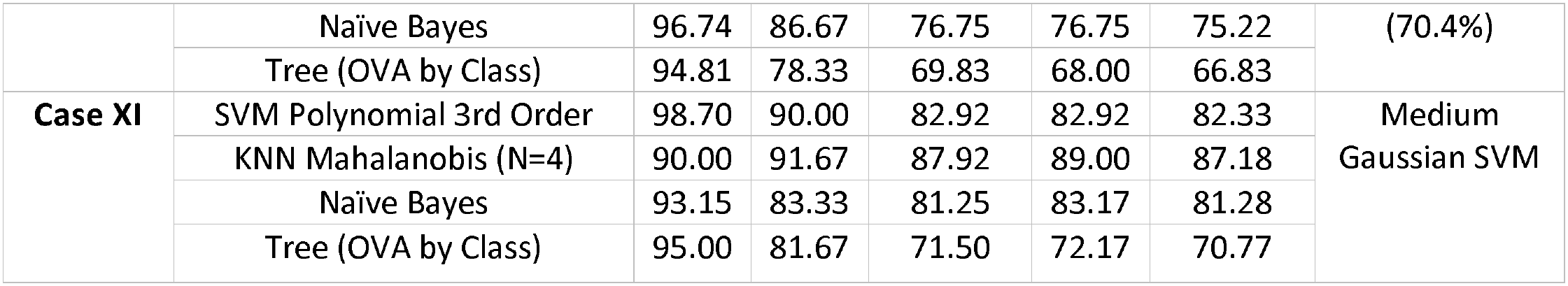
Classification performance of machine-learning models for motor imagery tasks relative to the resting baseline. Abbreviations: BEO, both eyes open; KNN, k-nearest neighbors; ML, machine learning; OVA, one-versus-all; SVM, support vector machine.

**Table 4.**
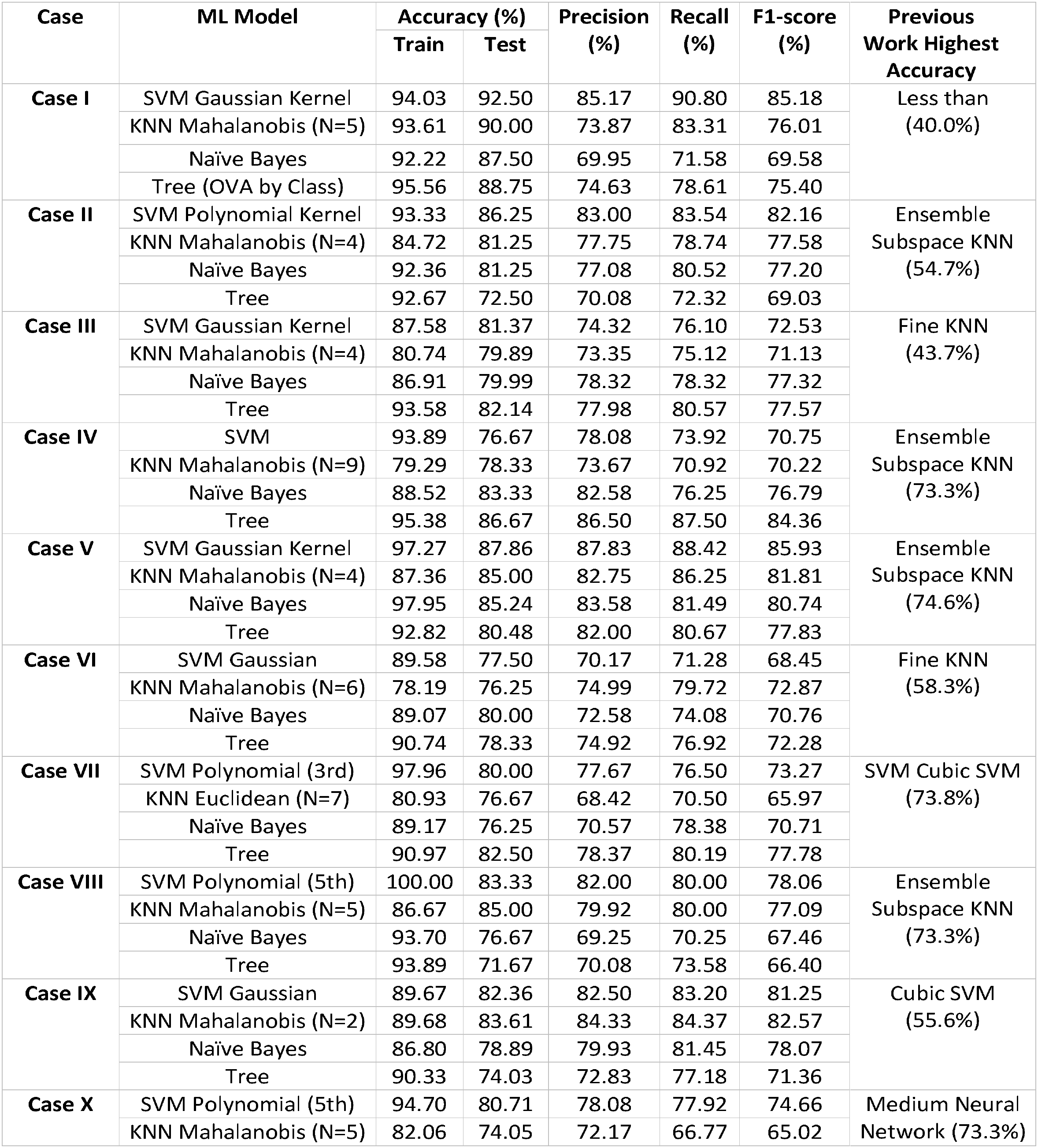
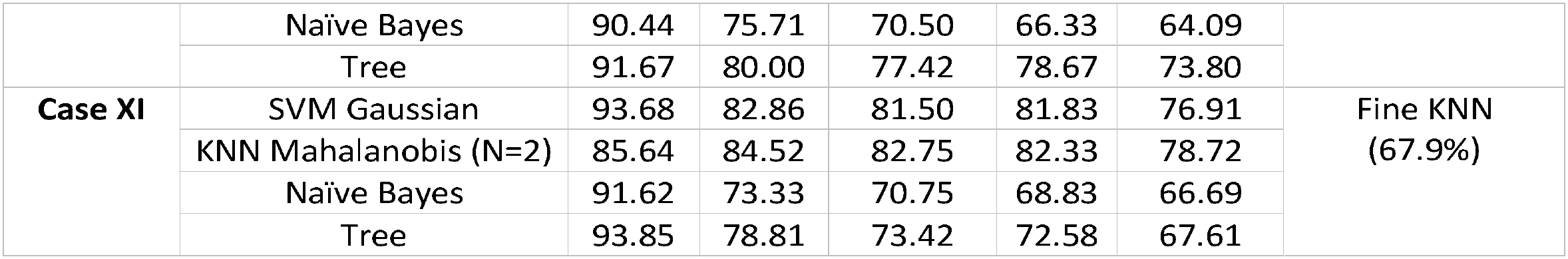
Classification performance of machine-learning models for motor execution tasks relative to the resting baseline. Abbreviations: BEO, both eyes open; KNN, k-nearest neighbors; ML, machine learning; OVA, one-versus-all; SVM, support vector machine.

### Motor Imagery Classification

Classification results for motor imagery tasks against the BEO baseline are presented in **Table 3**, with the distribution of best-performing classifiers across cases illustrated in **Figure 6**. In Case I, encompassing all six motor imagery movements, Gaussian-kernel SVM achieved the highest testing accuracy of 95.48%, with a training accuracy of 98.46%, precision of 86.97%, recall of 88.33%, and Fl-score of 87.04%. Na"ive Bayes achieved the second-highest testing accuracy (92.14%), followed by Mahalanobis KNN (90.71%) and decision tree (86.67%).

**Figure 6.**
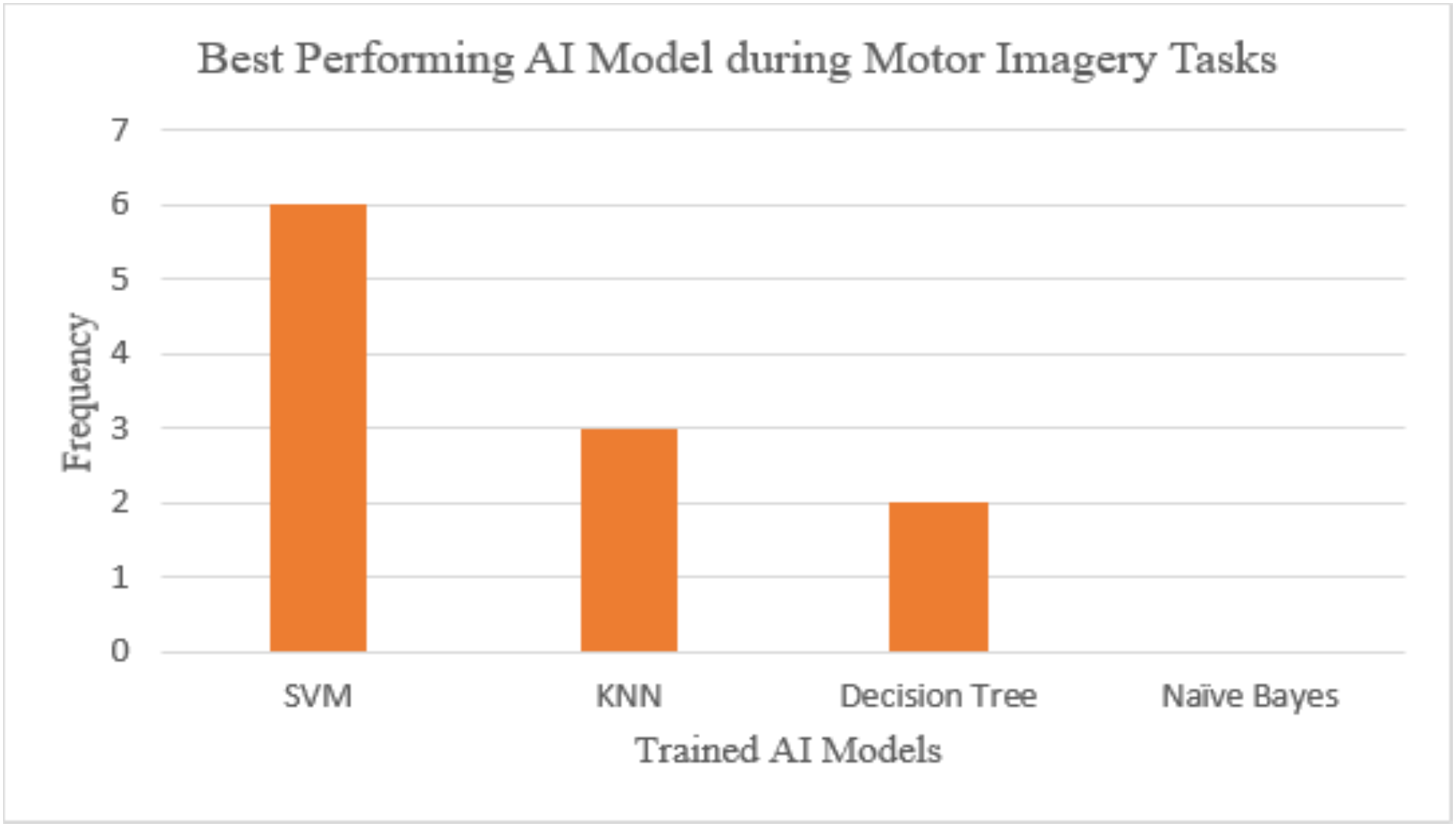
Frequency of best-performing machine-learning classifiers across motor imagery classification cases. Abbreviations: Al, artificial intelligence; KNN, k-nearest neighbors; SVM, support vector machine.

Performance varied across individual movement comparisons. Polynomial SVM achieved the highest testing accuracies in Cases II and IX (85.00% each), whereas Gaussian SVM and decision tree achieved identical accuracies of 86.67% in Case 111, with the decision tree producing a higher Fl-score (76.47% versus 73.39%). Mahalanobis KNN performed best in Cases X and XI, achieving testing accuracies of 88.33% and 91.67%, respectively. In Case XI, KNN also achieved an Fl-score of 87.18%, compared with 82.33% for polynomial SVM. Overall, Gaussian SVM provided the highest testing accuracy for the combined motor imagery comparison, while other classifiers demonstrated competitive performance in individual cases.

### Motor Execution Classification

Classification results for motor execution tasks against BEO are presented in **Table 4**, with the best-performing classifiers illustrated in **Figure 7**. In Case I, Gaussian-kernel SVM achieved the highest testing accuracy of 92.50%, with a training accuracy of 94.03%, precision of 85.17%, recall of 90.80%, and Fl-score of 85.18%. Mahalanobis KNN achieved a testing accuracy of 90.00%, followed by decision tree (88.75%) and Na"ive Bayes (87.50%).

**Figure 7.**
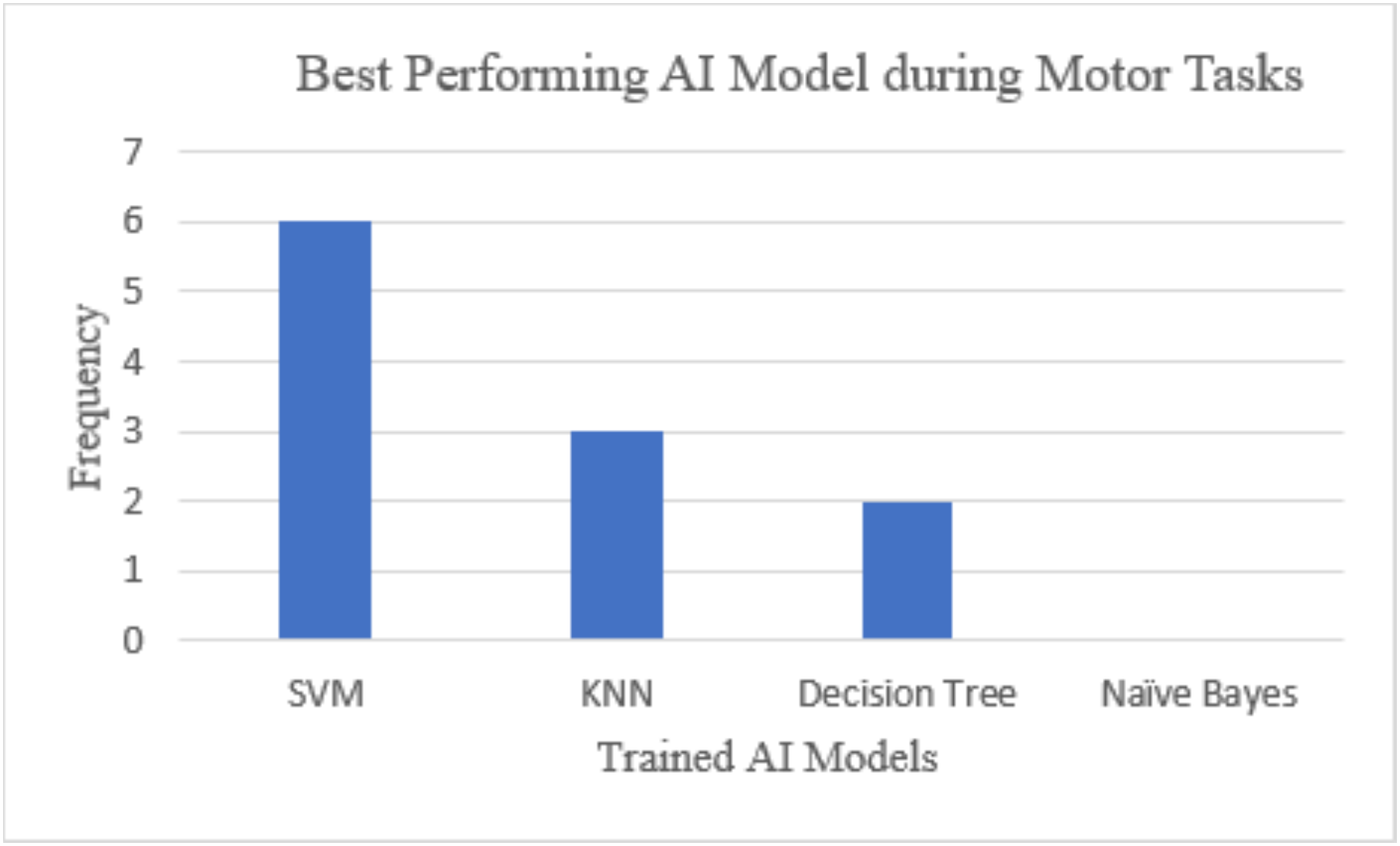
Frequency of best-performing machine-learning classifiers across motor execution classification cases. Abbreviations: Al, artificial intelligence; KNN, k-nearest neighbors; SVM, support vector machine.

For individual movement comparisons, polynomial SVM achieved the highest testing accuracy in Case II (86.25%; Fl-score: 82.16%). Decision tree performed best in Cases Ill and IV, with testing accuracies of 82.14% and 86.67%, respectively, while Gaussian SVM achieved the highest accuracy in Case V (87.86%; Fl-score: 85.93%). Mahalanobis KNN outperformed the other classifiers in Cases VIII, IX, and XI, achieving testing accuracies of 85.00%, 83.61%, and 84.52%, respectively. These findings demonstrate that although Gaussian SVM achieved the highest overall testing accuracy for the combined motor execution comparison, classifier performance varied across individual movementtasks.

### Comparison of Motor Imagery and Motor Execution

For the combined movement comparisons in Case I, Gaussian-kernel SVM achieved higher testing accuracy for motor imagery (95.48%) than motor execution (92.50%), representing a difference of 2.98 percentage points **(Tables 3 and 4).** The corresponding Fl-scores were 87.04% and 85.18%, respectively, representing a difference of 1.86 percentage points. However, the highest-performing classifier differed across individual movement comparisons, indicating that classification performance was task-dependent.

### Classification Performance Following Feature Selection

All reported classification results were obtained using four MRMR-selected features from the original 416-dimensional feature representation. Despite this substantial reduction in feature dimensionality, testing accuracies exceeding 90% were achieved for both combined movement comparisons. However, because classification performance using the complete feature set was not evaluated as a direct comparator, the results do not establish whether MRMR improved accuracy relative to the original feature representation

## Discussion

The present study investigated a multidomain feature-engineering framework for classifying motor execution and imagery signals using the MllimbEEG dataset. The principal finding was that strong classification performance could be achieved using only four MRMR-selected features from an initial set of 416 time-domain, frequency-domain, and wavelet-based features. Gaussian-kernel SVM achieved the highest overall testing accuracies of 92.50% for motor execution and 95.48% for motor imagery, with corresponding Fl-scores of 85.18% and 87.04%, respectively. These findings suggest that combining complementary EEG features with targeted feature selection can provide a compact representation for discriminating motor-related neural activity.

Classifier performance varied across individual movement comparisons. Although Gaussian SVM achieved the highest overall accuracies, Mahalanobis KNN and decision tree classifiers performed better in several specific cases. Greater difficulty was observed in distinguishing right-foot dorsiflexion imagery from baseline activity, potentially reflecting the subtle sensorimotor changes and low signal-to-noise ratio associated with Ml. These findings highlight the importance of evaluating task-specific classification performance rather than relying exclusively on aggregate accuracy.

The reduction from 416 features to four represents a substantial decrease in feature dimensionality while retaining high classification accuracy. By maximizing feature relevance and minimizing redundancy, MRMR provided a compact representation of the extracted EEG characteristics. Such dimensionality reduction may be advantageous for developing computationally efficient BCI systems, although direct comparisons of computational time and classification performance between the complete and reduced feature sets are needed to establish these benefits.

Previous studies have explored different strategies for improving Ml classification through channel and feature selection. Handiru et al. reported an average accuracy of approximately 63% using optimized EEG channel selection across 85 participants^18^. Xu et al. achieved a maximum accuracy of approximately 93% using ECoG-based classification with selected channels and combined features^19^. Similarly, Qiu et al. reported mean accuracies of 78% and 83% using an improved channel-selection approach across two BCI datasets2°. In comparison, the present study achieved 95.48% testing accuracy for motor imagery using four selected features from non-invasive EEG recordings. However, differences in recording modalities, datasets, classification tasks, and validation procedures prevent direct conclusions regarding comparative superiority.

Other investigations have emphasized dimensionality reduction and optimization for Ml classification. Mohammadi et al. reported approximately 64% mean accuracy using CSP and temporal and time-frequency features^21^. More recently, Olvera et al. achieved 83.82% mean accuracy using a quantum-based feature-selection approach with subject-dependent validation^22^. Unlike these approaches, the present study employed conventional MRMR feature selection with multidomain EEG features. The findings suggest that relatively straightforward feature-selection methods can also support strong classification performance, without necessarily requiring more complex optimization strategies.

An additional strength of this study is the evaluation of both upper- and lower-limb motor execution and imagery across 11 classification cases using recordings from multiple participants. This enabled assessment of classifier performance across different movement comparisons and demonstrated that classification accuracy was not uniform across tasks. However, several limitations should be acknowledged. Although 10-fold cross-validation was employed, the current methodology does not establish whether training and testing observations were separated by participant, limiting conclusions about subject-independent generalization. Furthermore, only a subset of the available MllimbEEG recordings was analyzed, and the complete 416-feature representation was not directly compared with the reduced feature set. The absence of external validation and real-time evaluation also limits conclusions regarding clinical or online BCI applicability.

Overall, the findings support the potential of multidomain EEG feature engineering combined with MRMR selection for motor execution and imagery classification. The substantial reduction in feature dimensionality, together with the observed classification performance, provides a basis for further investigation of compact EEG decoding frameworks. Future studies should incorporate subject-independent validation, larger datasets, direct computational comparisons, and real-time testing to establish the robustness and practical applicability of this approach in neurorehabilitation.

## Conclusion

This study demonstrated that multidomain EEG feature engineering combined with MRMR feature selection can support accurate classification of motor execution and motor imagery tasks using a substantially reduced feature representation. From 416 extracted features, four were selected for classification, with Gaussian-kernel SVM achieving the highest testing accuracies of 95.48% for motor imagery and 92.50% for motor execution. KNN and decision tree classifiers also demonstrated competitive performance across individual movement comparisons. These findings highlight the potential of compact EEG feature representations for developing less complex BCI systems for neurorehabilitation. However, further evaluation using subject-independent validation, larger datasets, and real-time testing is necessary to establish generalizability and practical applicability. Future work will focus on integrating the proposed framework with neuroprosthetic hardware and evaluating its performance in real-world rehabilitation settings.

## Data Availability

Data used in this research work is available at: MILimbEEG: An EEG Signals Dataset based on Upper and Lower Limb Task During the Execution of Motor and Motor imagery Tasks - Mendeley Data

https://doi.org/10.17632/x8psbz3f6x.2

## Disclosures

### Author contributions

SSA: conceptualization, writing-original draft, methodology, investigation, formal analysis.

SJ: methodology, formal analysis, conceptualization, writing-review and editing.

MAK: writing-original draft, methodology, investigation.

ZZF: investigation, writing-review and editing.

TS: supervision, writing-original draft, methodology.

MB: writing-original draft, writing-review and editing.

RAK: formal analysis, writing-review and editing.

### Data statement

Data used in this research work is available at: MllimbEEG: An EEG Signals Dataset based on Upper and Lower Limb Task During the Execution of Motor and Motor imagery Tasks-Mendeley Data

## Appendix

## Abbreviation

AAM: Adaptive Autoregressive Modelling
AES: American EEG Society
AI: Artificial Intelligence
BCI: Brain–Computer Interface
BEO: Both Eyes Open
CLH: Closing Left Hand
CRH: Closing Right Hand
CSP: Common Spatial Pattern
CSV: Comma-Separated Values
DFT: Discrete Fourier Transform
DLF: Dorsal Flexion of Left Foot
DRF: Dorsal Flexion of Right Foot
DWT: Discrete Wavelet Transform
ECoG: Electrocorticography
EEG: Electroencephalography
ERP: Event-Related Potential
ESPOL: Escuela Superior Politécnica del Litoral
FFT: Fast Fourier Transform
FN: False Negative
FP: False Positive
HA: Hjorth Activity
HC: Hjorth Complexity
HM: Hjorth Mobility
ICP: Intrahepatic Cholestasis of Pregnancy
IMOCS: Iterative Multi-objective Optimization for Channel Selection
KNN: K-Nearest Neighbors
LARS: Least-Angle Regression
LDA: Linear Discriminant Analysis
LSL: Lab Streaming Layer
ME: Mean Energy
MI: Motor Imagery
ML: Machine Learning
MRMR: Maximum Relevance Minimum Redundancy
MTE: Mean Teager’s Energy
NFD: Normalized First Difference
NQGA: Novel Genetic Quantum Algorithm
NSD: Normalized Second Difference
OLS: Ordinary Least Squares
OVA: One-vs-All
PCA: Principal Component Analysis
PLF: Plantar Flexion of Left Foot
PRF: Plantar Flexion of Right Foot
PSD: Power Spectral Density
SCP: Slow Cortical Potential
SD: Standard Deviation
SFFS: Sequential Floating Forward Selection
ShEn: Shannon Entropy
SSVEP: Steady-State Visual Evoked Potential
SVM: Support Vector Machine

## References

1. Global report on health equity for persons with disabilities. Accessed October 13, 2026. https://www.who.int/publications/i/item/9789240063600

2. Wolpaw JR, Millan JDR, Ramsey NF. Brain-computer interfaces: Definitions and principles. Handb Clin Neurol. 2020;168:15–23. doi:10.1016/B978-0-444-63934-9.00002-0

3. Khan MA, Das R, Iversen HK, Puthusserypady S. Review on motor imagery based BCI systems for upper limb post-stroke neurorehabilitation: From designing to application. Comput Biol Med. 2020;123:103843. doi:10.1016/j.compbiomed.2020.103843

4. Neuper C, Scherer R, Wriessnegger S, Pfurtscheller G. Motor imagery and action observation: modulation of sensorimotor brain rhythms during mental control of a brain-computer interface. Clin Neurophysiol Off 1 Int Fed Clin Neurophysiol. 2009;120(2):239–247. doi:10.1016/j.clinph.2008.11.015

5. Khademi Z, Ebrahimi F, Kordy HM. A review of critical challenges in MI-BCI: From conventional to deep learning methods. J Neurosci Methods. 2023;383:109736. doi:10.1016/j.jneumeth.2022.109736

6. Schirrmeister RT, Springenberg JT, Fiederer LDJ, et al. Deep learning with convolution al neural networks for EEG decoding and visualization. Hum Brain Mapp. 2017;38(11):5391–5420. doi:10.1002/hbm.23730

7. Al-Saegh A, Dawwd SA, Abdul-Jabbar JM. Deep learning for motor imagery EEG-based classification: A review. Biomed Signal Process Control. 2021;63:102172. doi:10.1016/j.bspc.2020.102172

8. Padfield N, Zabalza J, Zhao H, Masero V, Ren J. EEG-Based Brain-Computer Interfaces Using Motor-Imagery: Techniques and Challenges. Sensors. 2019;19(6):1423. doi:10.3390/s19061423

9. Kumar S, Sharma A, Tsunoda T. An improved discriminative filter bank selection approach for motor imagery EEG signal classification using mutual information. BMC Bioinformatics. 2017;18(Suppl 16):545. doi:10.1186/s12859-017-1964-6

10. Tangermann M, Muller KR, Aertsen A, et al. Review of the BCI Competition IV. Front Neurosci. 2012;6:55. doi:10.3389/fnins.2012.00055

11. Gwon D, Won K, Song M, Nam CS, Jun SC, Ahn M. Review of public motor imagery and execution datasets in brain-computer interfaces. Front Hum Neurosci. 2023;17:1134869. doi:10.3389/fnhum.2023.1134869

12. Asanza V, Montoya D, Lorente-Leyva LL, Peluffo-Ordonez DH, Gonzalez K. MILimbEEG: An EEG Signals Dataset based on Upper and Lower Limb Task During the Execution of Motor and Motorimagery Tasks. 2023;2. doi:10.17632/x8psbz3f6x.2

13. Asanza V, Lorente-Leyva LL, Peluffo-Ordonez DH, Montoya D, Gonzalez K. MILimbEEG: A dataset of EEG signals related to upper and lower limb execution of motor and motor imagery tasks. Data Brief. 2023;50:109540. doi:10.1016/j.dib.2023.109540

14. Welch P. The use of fast Fourier transform for the estimation of power spectra: A method based on time averaging over short, modified periodograms. IEEE Trans Audio Electroacoustics. 1967;15(2):70–73. doi:10.1109/TAU.1967.1161901

15. Alturki FA, AISharabi K, Abdurraqeeb AM, Aljalal M. EEG Signal Analysis for Diagnosing Neurological Disorders Using Discrete Wavelet Transform and Intelligent Techniques. Sensors. 2020;20(9):2505. doi:10.3390/s20092505

16. Jawed S, Amin HU, Malik AS, Faye I. Classification of Visual and Non-visual Learners Using Electroencephalographic Alpha and Gamma Activities. Front Behav Neurosci. 2019;13:86. doi:10.3389/fnbeh.2019.00086

17. Hawkins DM, Basak SC, Mills D. Assessing model fit by cross-validation.J Chem Inf Comput Sci. 2003;43(2):579–586. doi:10.1021/ci025626i

18. Handiru VS, Prasad VA. Optimized Bi-Objective EEG Channel Selection and Cross-Subject Generalization with Brain-Computer Interfaces. IEEE Trans Hum-Mach Syst. 2016;46(6):777–786. doi:10.1109/THMS.2016.2573827

19. Classification of motor imagery tasks for electrocorticogram based brain-computer interface | Biomedical Engineering Letters | Springer Nature Link. Accessed October 13, 2026. https://link.springer.com/article/10.1007/s13534-014-0128-0

20. Qiu Z, Jin J, Lam HK, Zhang Y, Wang X, Cichocki A. Improved SFFS method for channel selection in motor imagery based BCI. Neurocomputing. 2016;207(C):519–527. doi:10.1016/j.neucom.2016.05.035

21. Mohammadi E, Daneshmand PG, Khorzooghi SMSM. Electroencephalography-Based Brain-Computer Interface Motor Imagery Classification. J Med Signals Sens. 2022;12(1):40–47. doi:10.4103/jmss.JMSS_74_20

22. Olvera C, Ross OM, Rubio Y. EEG-based motor imagery classification with quantum algorithms. Expert Syst Appl. 2024;247:123354. doi:10.1016/j.eswa.2024.123354

